# Impacts of Social and Economic Factors on the Transmission of Coronavirus Disease 2019 (COVID-19) in China

**DOI:** 10.1101/2020.03.13.20035238

**Authors:** Yun Qiu, Xi Chen, Wei Shi

## Abstract

This paper examines the role of various socioeconomic factors in mediating the local and cross-city transmissions of the novel coronavirus 2019 (COVID-19) in China. We implement a machine learning approach to select instrumental variables that strongly predict virus transmission among the rich exogenous weather characteristics. Our 2SLS estimates show that the stringent quarantine, massive lockdown and other public health measures imposed in late January significantly reduced the transmission rate of COVID-19. By early February, the virus spread had been contained. While many socioeconomic factors mediate the virus spread, a robust government response since late January played a determinant role in the containment of the virus. We also demonstrate that the actual population flow from the outbreak source poses a higher risk to the destination than other factors such as geographic proximity and similarity in economic conditions. The results have rich implications for ongoing global efforts in containment of COVID-19.

## 1 Introduction

Several clusters of patients with pneumonia of unknown cause were reported in late December 2019 in Wuhan, the capital city of Hubei Province, China. It was later identified to be caused by a new coronavirus (severe acute respiratory syndrome coronavirus 2, or SARS-CoV-2) (Zhu *et al*., 2020) and the disease is named by the World Health Organization (WHO) as Coronavirus Disease 2019 (COVID-19)^1^. Since mid January 2020, it has rapidly spread throughout China and other countries. The first confirmed case outside Wuhan in China was reported on January 19 (Shenzhen) (Li *et al*., 2020). Similar to SARS-CoV and MERS-CoV, the COVID-19 can be transmitted from person to person. As of January 30, a total of 9976 cases had been reported in at least 21 countries^2^. Early detection of COVID-19 importation and prevention of onward transmission are crucial to all areas at risk of importation from areas with active transmissions (Gilbert *et al*., 2020). To contain the virus, Wuhan was placed under lockdown with residents not allowed to leave the city on January 23. On the same or the next day, most public transportation stopped in Wuhan and other cities in Hubei province. In addition, most cities in Hubei province had adopted blockade measures for Class A infectious diseases by January 28^3^. Residents in those areas are highly encouraged to stay at home and not to attend any gathering activity. Besides, at least 207 cities in China have adopted similar though softer measures by February 12. People are frequently reminded by media and community workers to maintain safe social distance. Furthermore, many cities require that recent visitors to high COVID-19 risk areas quarantine themselves at home or in designated facilities for 14 days. Residents who have symptoms of fever or dry cough are required to report the situation to community, and are quarantined and treated in special medical facilities. Governments also trace and isolate contacts of those patients.

As multiple nations have implemented mandatary quarantine or even massive lockdown, such as South Korea, Italy and China, and more countries are expected to join as the coronavirus outbreak becomes a global pandemic, examining the influencing factors of the transmission of COVID-19 and the effectiveness of the large-scale quarantine measures in China not only adds to our understanding of the containment of COVID-19 but also provides insights into future prevention work against similar infectious diseases. In this paper, we estimate how the number of confirmed COVID-19 cases in a city is influenced by the number of new COVID-19 cases in the same city, other cities, and Wuhan in the preceding first and second weeks, respectively. China’s large scale quarantine policy takes effect in late January. We also test whether this policy succeeds in delaying the spread of COVID-19 using a 14-day rolling window analysis between January 19 and February 15.

For infectious diseases, the number of infected people usually increases first before reaching a peak and then drops. This pattern implies that for a linear equation of new cases on the number of cases in the past, the unobserved determinants of new infections are serially correlated. The unobserved determinants of new cases may be serially correlated, also because they measure persistent factors, such as people’s habit and government policy. Serial correlations in errors give rise to correlations between the lagged number of cases and the error term, and ordinary least square (OLS) estimator may be biased. Combining insights in Adda (2016) and the existing knowledge of the incubation period of COVID-19, we construct instrumental variables for the number of new COVID-19 cases in the preceding two weeks. Weather characteristics in the previous third and fourth weeks do not directly affect today’s number of new COVID-19 cases after controlling for the number of new COVID-19 cases and weather conditions in the preceding first and second weeks. Therefore, our estimated impacts have causal interpretations. We use the Lasso method to select instrumental variables among eleven weather characteristics that have the highest predictive power for the average number of new confirmed cases during each of the past two weeks. Furthermore, we examine the moderator effects of socioeconomic factors on the transmission of COVID-19 in China, which include population flow out of Wuhan, the distance between cities, GDP per capita, number of doctors, and contemporaneous weather characteristics. We focus on the effect of population flows from the origin of the COVID-19 outbreak, because data on real time travel between cities have recently become available and we examine whether it can explain the disease spread, and that Wuhan is a major city and a transportation hub with significant population movements.

We find that the stringent quarantine and public health measures imposed in late January significantly decreased the transmission rate of COVID-19. By early February, the spread of the virus had been contained in China. While many socioeconomic factors moderate the spread of the virus, the actual population flow from the source poses a higher risk to the destination than other factors such as geographic proximity and similarity in economic conditions.

Our analysis contributes to the existing literature in two aspects. First, our analysis is connected to the economics and epidemiologic literature on the influencing factors of and ways of preventing the spread of infectious diseases. Existing studies find that reductions in interpersonal contact from holiday school closings (Adda, 2016), reactive school closure (Litvinova *et al*., 2019), public transportation strikes (Godzinski and Suarez Castillo, 2019), strategic targeting of travelers from high-incidence locations (Milusheva, 2017), and paid sick leave to keep contagious workers at home (Barmby and Larguem, 2009, Pichler and Ziebarth, 2017) can reduce the prevalence of influenza. Additionally, literature find that epidemics spread faster during economic booms (Adda, 2016), increases in employment are associated with increased incidence of influenza (Markowitz *et al*., 2010), and growth in trade can significantly increase the spread of influenza (Adda, 2016) and HIV (Oster, 2012). Vaccination (Maurer, 2009, White, 2019) and sunlight exposure (Slusky and Zeckhauser, 2018) are also found effective in reducing the spread of influenza. In the case of COVID-19, the quarantine policy takes effect throughout China within few days in late January. Thus, we directly compare the transmission effects in the pre and post February 1 subsamples to examine its effectiveness in preventing the transmission of COVID-19. Interestingly, within one week, population flow from Wuhan significantly increased the spread of virus in the early subsample, but decreased its transmission rate in the later subsample, suggesting that people have been taking more cautious measures from high COVID-19 risk areas. Besides, following the previous literature on the moderator effect of economic development and environmental characteristics on virus transmission, our analysis further reveal that COVID-19 transmission is positively moderated by GDP per capita and negatively moderated by the number of doctors, while the effects of the environmental factors and population density are mixed.

Second, our paper complements the epidemiologic studies on the basic reproduction number of COVID-19. Using data on the first 425 COVID-19 patients by January 22, Li *et al*. (2020) estimate a basic reproduction number of 2.2. Based on time-series data on the number of COVID-19 cases in mainland China from January 10 to January 24, Zhao *et al*. (2020) estimate that the mean reproduction number ranges from 2.24 to 3.58. Given the short time since the beginning of the COVID-19 outbreak, more research in this area is required to assess the dynamics of transmission and its implications on how the COVID-19 outbreak will evolve (Wu and McGoogan, 2020, Wu *et al*., 2020b). Several recent studies have examined the effect of population movement on the spread of COVID-19 (Zhan *et al*., 2020, Zhang *et al*., 2020). Our analysis relies on spatially disaggregated data in a longer period, and the instrumental variable approach we use imposes fewer restrictions on the relationship between the unobserved determinants of new cases and the number of cases in the past. Using exogenous temperature, wind speed and precipitation in the preceding third and fourth weeks as the instruments, we find that in the January 19 - February 1 subsample, one new infection leads to 1.465 more cases within one week and the effect on the second week is not definitive. While this causal estimate is slightly lower than the above epidemiologic studies, the rolling window analysis shows that the infection rate increases first in late January, then stabilizes and drops. Our paper also adds to the epidemiologic studies on effective ways of containing COVID-19. Using a stochastic transmission model that is parameterized to the COVID-19 outbreak, Hellewell *et al*. (2020) find that highly effective contact tracing and case isolation is enough to control a new outbreak of COVID-19 within three months in most scenarios. Our empirical estimation based on real numbers of newly confirmed COVID-19 cases suggests that one new infection case can decrease the number of new cases by 0.097 (0.438) in the following first (second) week in the February 2-15 subsample, corroborating the effectiveness of China’s large-scale quarantine policy.

This paper is organized as follows. Section 2 introduces the empirical model. Section 3 discusses our data and the construction of key variables. The results are presented in Section 4. Section 5 concludes.

## 2 Empirical Model

Our analysis sample includes 303 prefecture-level cities in China. We exclude Wuhan from our analysis because the epidemic patterns in Wuhan are significantly different from those in other cities. Some confirmed cases in Wuhan contracted the virus through exposure to Huanan Seafood Wholesale Market, which is the most probable origin of the virus^4^. In other cities, infections arise from human to human transmissions. The health care system in Wuhan faced the challenge of previously unknown virus infections in early January and became overwhelmed as the number of new cases increased exponentially from mid-January. These factors may cause severe delay and measurement error issues in the number of cases reported in Wuhan. We alleviate the measurement error problem using variations in the infections caused by exogenous changes in weather conditions which is discussed in detail in Section 3.2.

To model the spread of the virus, we consider simultaneously within city spread and between city transmissions, as in Adda (2016). The baseline model is

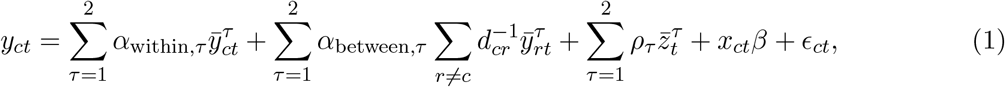

where *c* is a city other than Wuhan, *y*_*ct*_ is the number of new confirmed cases of COVID-19 in city *c* and date *t. τ* denotes the preceding first or second week. *d*_*cr*_ is the distance between cities *c* and *r* in log, and 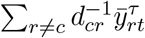 is the inverse distance weighted average of new infections in other cities. We include lagged *y*_*ct*_ up to 14 days based on the estimates of the durations of the infectious period and the incubation period in the literature^5^. We take average of lagged *y*_*ct*_ by week, as 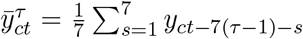 and 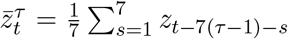. In addition, a notable feature of the COVID-19 epidemic is that it was originated in one city (Wuhan) and most of the early cases outside Wuhan can be traced to previous contacts with persons in Wuhan. To model how the virus spreads to other cities from its source, we include the number of new confirmed cases in Wuhan in the model 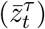 *x*_*ct*_ includes weather controls, city, and day fixed effects^6^. *Є*_*ct*_ is the error term. Standard errors are clustered within provinces. In a simplified model, we consider only within city transmissions,

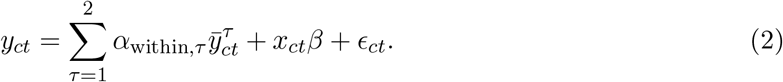

There are several reasons that 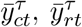 and 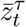 may be correlated with the error term *Є*_*ct*_. The unobserved determinants of new infections such as local residents’ and government’s preparedness are likely correlated over time, which causes correlations between the error term and the lagged dependent variables. Therefore, the coefficients are estimated by two-stage least squares in order to obtain consistent estimates. For the *y*_*ct*_ equation (2), the instrumental variables include average temperature, maximum wind speed and precipitation for city *c* for the past three and four weeks. The selection of weather variables as instruments is discussed in detail in Section 3.2. The timeline of key variables are displayed in Appendix Figure A.1. The primary assumption on the instrumental variables is that weather conditions before two weeks do not affect the likelihood that a person susceptible to the virus contracts the disease, conditional on weather conditions within two weeks. On the other hand, they affect the number of other persons who have become infectious within the two-week window, because they may have contracted the virus earlier than two weeks. These weather variables are exogenous to the error term and affect the spread of the virus, and have been used by Adda (2016) to instrument flu infections^7^.

A main objective of this paper is to quantify the effect of various social and economic factors in mediating the transmission rates of the virus, which may help identify potential behavioral and socioeconomic risk factors for infections. For within city transmissions, we consider the mediating effect of population density, degree of economic development, number of doctors, and environmental factors such as temperature, wind and rain. To measure the spread of the virus from Wuhan, we include an estimate of the number of people traveling from Wuhan. The empirical model including these mediating variables is as follows,

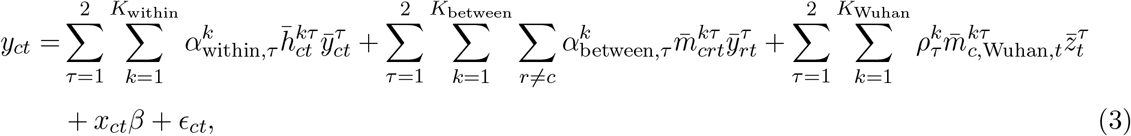

Where 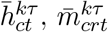 and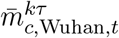 are the mediating factor for within city transmissions, between city transmissions and imported cases from Wuhan.

## 3 Data

### 3.1 Variables

January 19, 2020 is the first day that COVID-19 cases were reported outside of Wuhan, so we collect the daily number of new cases of COVID-19 for 303 cities from January 19 to February 15. All these data are reported by 32 provincial Health Commissions in China^8^. Figure 1 shows the time patterns of daily confirmed new cases in Wuhan, in Hubei province outside Wuhan, and in mainland China but outside Hubei province.

**Figure 1:**
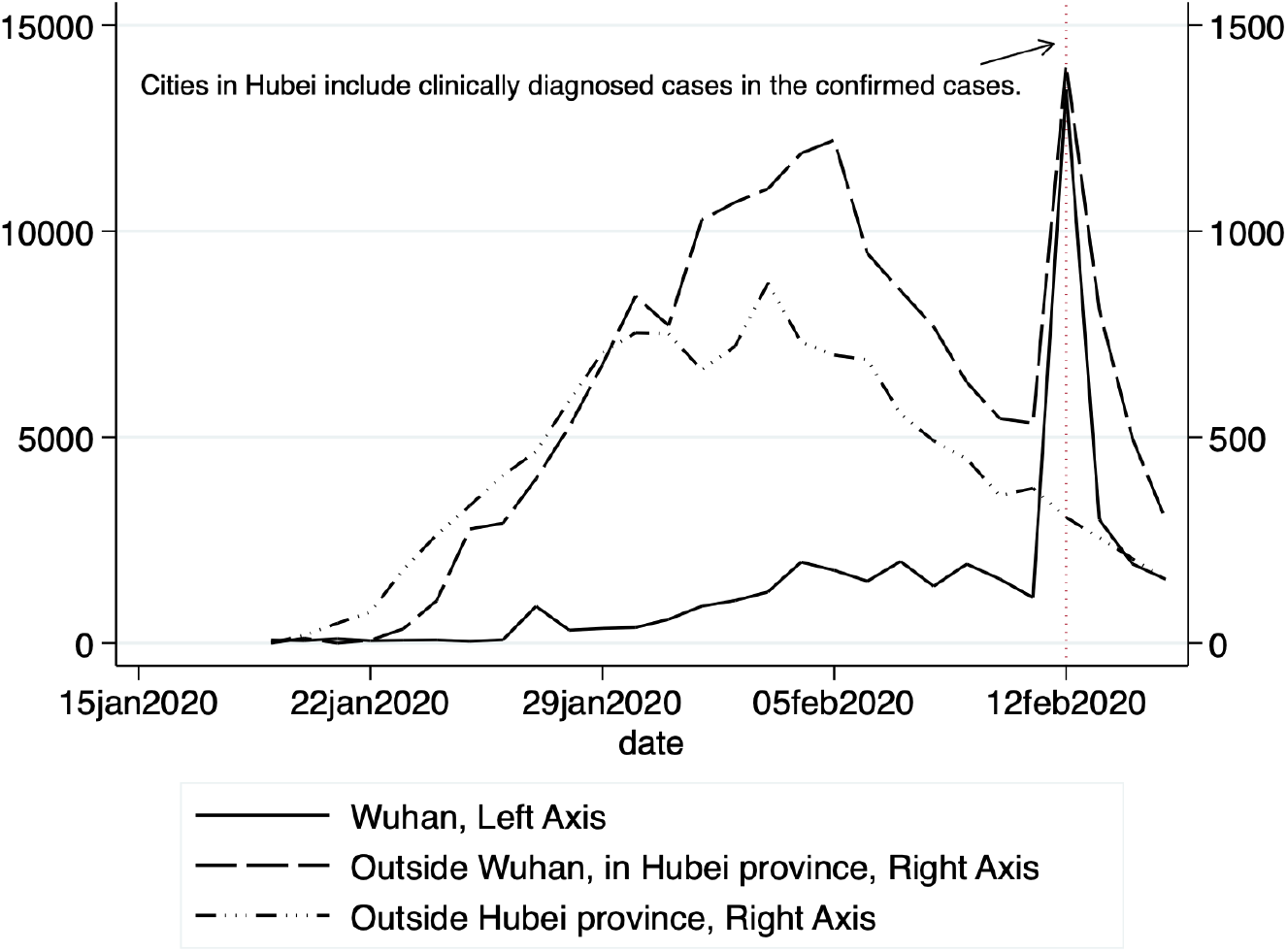
Number of Daily New Confirmed Cases of COVID-19 in Mainland China

For the explanatory variables, we calculate the number of new cases of COVID-19 in the preceding first and second weeks for each city on each day. To estimate the impacts of new COVID-19 cases in other cities on a city’s own COVID-19 new case, we first calculate the geographic distance between a city and all other cities using the latitudes and longitudes of the centroids of each city, and then calculate the weighted sum of the number of COVID-19 new cases in all other cities using the inverse of log distance between a city and each of the other cities as the weight.

Since the COVID-19 outbreak starts from Wuhan, we also calculate the weighted number of COVID-19 new cases in Wuhan using the inverse of log distance as the weight. Furthermore, to explore the mediating impact of population flow from Wuhan, we collect the daily population flow index from Baidu that proxies for the total intensity of migration from Wuhan to other cities^9^. Figure 2 plots the Baidu index of population flow out of Wuhan and compares its values this year with those in 2019. We then interact with the flow index the share that a destination city takes (Figure 3) to construct a measure on the population flow from Wuhan to a destination city. Other mediating variables include population density, GDP per capita, and the number of doctors at the city level, which we collect from the most recent China city statistical yearbook. Table 1 presents the summary statistics of these variables.

**Table 1:** Summary Statistics

| Variable | N | Mean | Std dev. | Min. | Median | Max. |
| --- | --- | --- | --- | --- | --- | --- |
| *Non Hubei cities* |  |  |  |  |  |  |
| City characteristics |  |  |  |  |  |  |
| GDP per capita, 10,000RMB | 287 | 5.236 | 3.024 | 1.141 | 4.334 | 21.549 |
| Population density, per km<sup>2</sup> | 287 | 430.247 | 374.071 | 9.049 | 332.180 | 3444.092 |
| # of doctors, 10,000 | 287 | 1.089 | 1.139 | 0.030 | 0.808 | 10.938 |
| Time varying variables, Jan 19 - Feb 1 |  |  |  |  |  |  |
| Daily # of new confirmed cases | 4018 | 1.307 | 3.614 | 0 | 0 | 60 |
| Weekly average temperature, °C | 4018 | 3.287 | 9.182 | -29.825 | 3.862 | 23.099 |
| Weekly average maximum wind speed, m/s | 4018 | 3.728 | 1.274 | 1.095 | 3.520 | 10.392 |
| Weekly average precipitation, mm | 4018 | 0.239 | 0.559 | 0 | 0.033 | 5.570 |
| Time varying variables, Feb 1 - Feb 15 |  |  |  |  |  |  |
| Daily # of new confirmed cases | 4018 | 1.708 | 3.539 | 0 | 0 | 49 |
| Weekly average temperature, °C | 4018 | 4.339 | 8.865 | -29.881 | 5.881 | 23.340 |
| Weekly average maximum wind speed, m/s | 4018 | 3.889 | 1.310 | 1.227 | 3.777 | 9.789 |
| Weekly average precipitation, mm | 4018 | 0.175 | 0.493 | 0.000 | 0.022 | 5.432 |
| *Cities in Hubei province, excluding Wuhan* |  |  |  |  |  |  |
| City characteristics |  |  |  |  |  |  |
| GDP per capita, 10,000RMB | 16 | 4.932 | 1.990 | 2.389 | 4.306 | 8.998 |
| Population density, per km<sup>2</sup> | 16 | 416.501 | 220.834 | 24.409 | 438.820 | 846.263 |
| # of doctors, 10,000 | 16 | 0.698 | 0.436 | 0.017 | 0.702 | 1.393 |
| Time varying variables, Jan 19 - Feb 1 |  |  |  |  |  |  |
| Daily # of new confirmed cases | 224 | 22.165 | 35.555 | 0 | 7 | 276 |
| Weekly average temperature, °C | 224 | 4.450 | 1.674 | -1.849 | 4.651 | 7.950 |
| Weekly average maximum wind speed, m/s | 224 | 3.179 | 0.888 | 1.178 | 3.066 | 6.026 |
| Weekly average precipitation, mm | 224 | 0.261 | 0.313 | 0.000 | 0.160 | 1.633 |
| Time varying variables, Feb 1 - Feb 15 |  |  |  |  |  |  |
| Daily # of new confirmed cases | 224 | 53.013 | 63.654 | 0 | 33.5 | 424 |
| Weekly average temperature, °C | 224 | 7.052 | 1.937 | -0.952 | 7.264 | 11.634 |
| Weekly average maximum wind speed, m/s | 224 | 3.237 | 1.018 | 1.132 | 3.184 | 6.827 |
| Weekly average precipitation, mm | 224 | 0.156 | 0.224 | 0.000 | 0.094 | 1.306 |

Variables of the city characteristics are obtained from City Statistical Yearbooks. Time varying variables are observed daily for each city. Weekly average weather variables are averages over the proceeding week.

**Figure 2:**
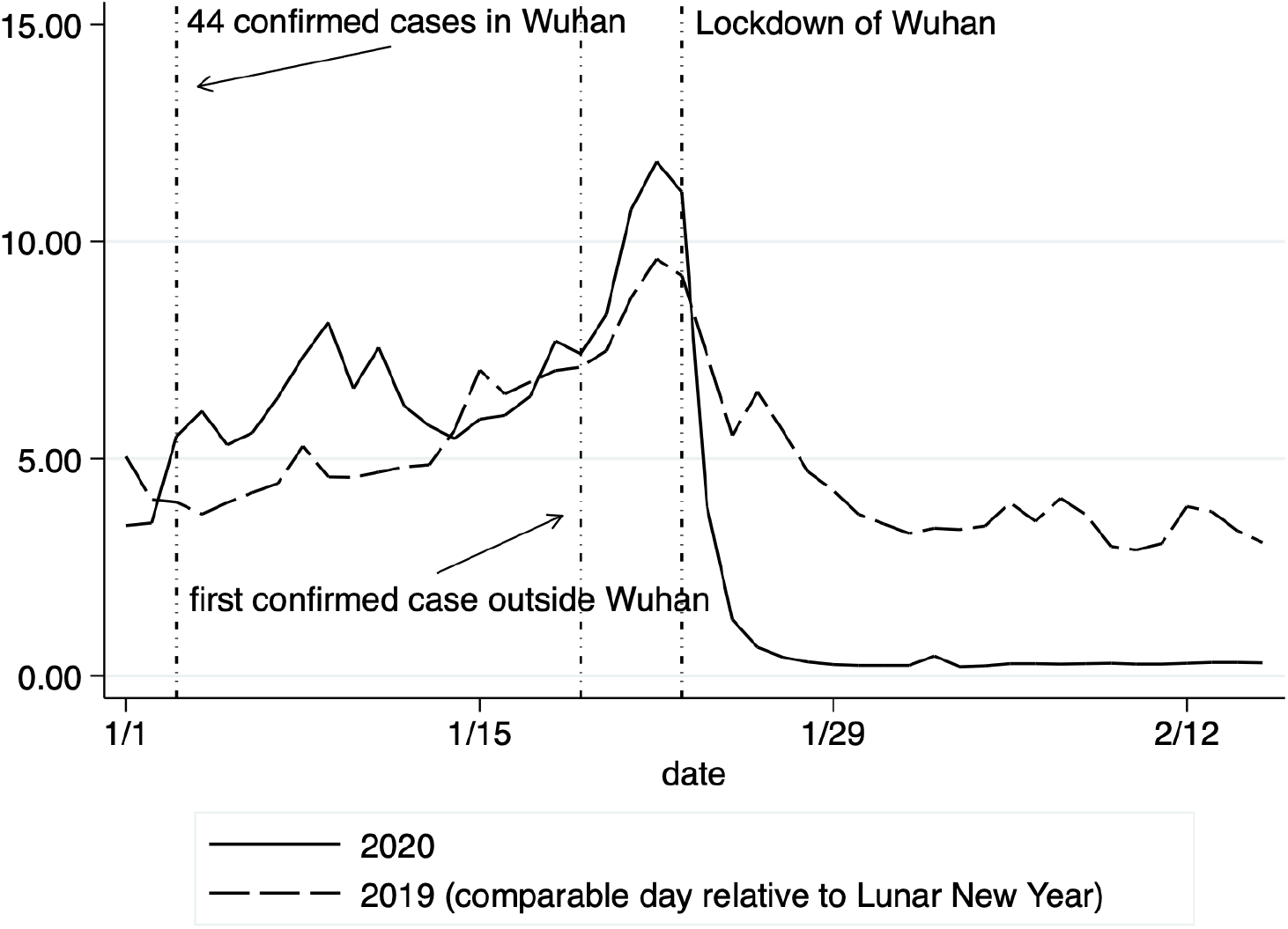
Baidu Index of Population Flow from Wuhan

**Figure 3:**
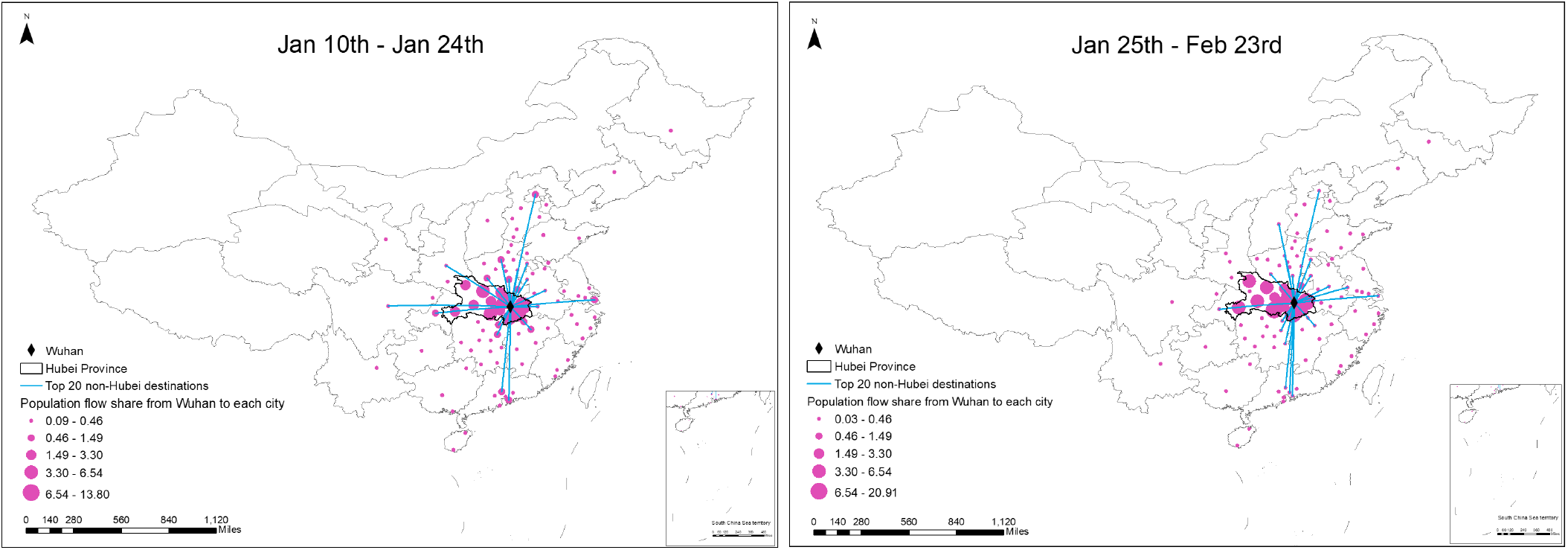
Destination Share in Population Flow from Wuhan

We rely on meteorological data to construct instrumental variables for the endogenous variables. The National Oceanic and Atmospheric Administration (NOAA) provides average, maximum and minimum temperatures, air pressure, average and maximum wind speeds, precipitation, snowfall amount, and dew point for 362 weather stations at the daily level in China. To merge the meteorological variables with the number of new cases of COVID-19, we first calculate daily weather variables for each city on each day from 2019 December to 2020 February from station-level weather records following the inverse-distance weighting method. Specifically, for each city, we draw a circle of 100 km from the city’s centroid and calculate the weighted average daily weather variables using stations within the 100 km circle^10^. We use the inverse of the distance between the city’s centroid and each station as the weight. Second, we match the daily weather variables to the number of new cases of COVID-19 based on city name and date.

### 3.2 Selection of Instrumental Variables

The transmission rate of COVID-19 may be affected by many environmental factors. Human-to-human transmission of COVID-19 is mostly through droplets and contacts (National Health Commission of the PRC, 2020). Weather conditions such as rainfall, wind speed, and temperatures may play a key role in shaping infections via influencing social activities and virus transmissions. For instance, more precipitation increases humidity that may weaken virus transmissions (Lowen and Steel, 2014). Both more rainfall and lower temperature may reduce social activities. Lower temperatures may also enhance the infectiousness of the virus in the environment (Wang *et al*., 2020). Higher wind speed and therefore ventilated air may disturb virus transmissions. Current new cases arise from contracting the virus in the past, and for the case of COVID-19, typically within two weeks. The extent of human-to-human transmission is determined by the number of people who already contracted the virus and the environmental conditions within two weeks. Conditional on the number of confirmed cases and environmental conditions within two weeks, it is plausible that weather conditions further in the past should not affect current new cases.

There are many potentially relevant weather characteristics, including daily average, maximum and minimum temperature, average and maximum wind speed, precipitation, air pressure, visibility, dew point, snow depth, and the presence of extreme weather. We use Lasso method to select environmental variables that have the highest predictive power for the average number of daily new confirmed cases during each of the past two weeks, with provincial and day fixed effects already included.The penalty parameters are given by the plugin method of Belloni *et al*. (2014). There are four variables selected, including average maximum wind speed and average precipitation during the past third and fourth weeks.We also include average temperature during the past third and fourth weeks because temperature is known to affect the transmissions of flu virus and is conjectured to affect COVID-19 as well (Wang *et al*., 2020). We then regress the endogenous variables on the one to four week lags of the instrumental variables, city, and date fixed effects. Table 2 shows that F-tests on the coefficients of the three and four week lags of the instrumental variables all strongly reject joint insignificance, which confirms that the selected instrumental variables are not weak. The coefficients of the first stage regressions are reported in Table B.1 in the appendix.

**Table 2:**
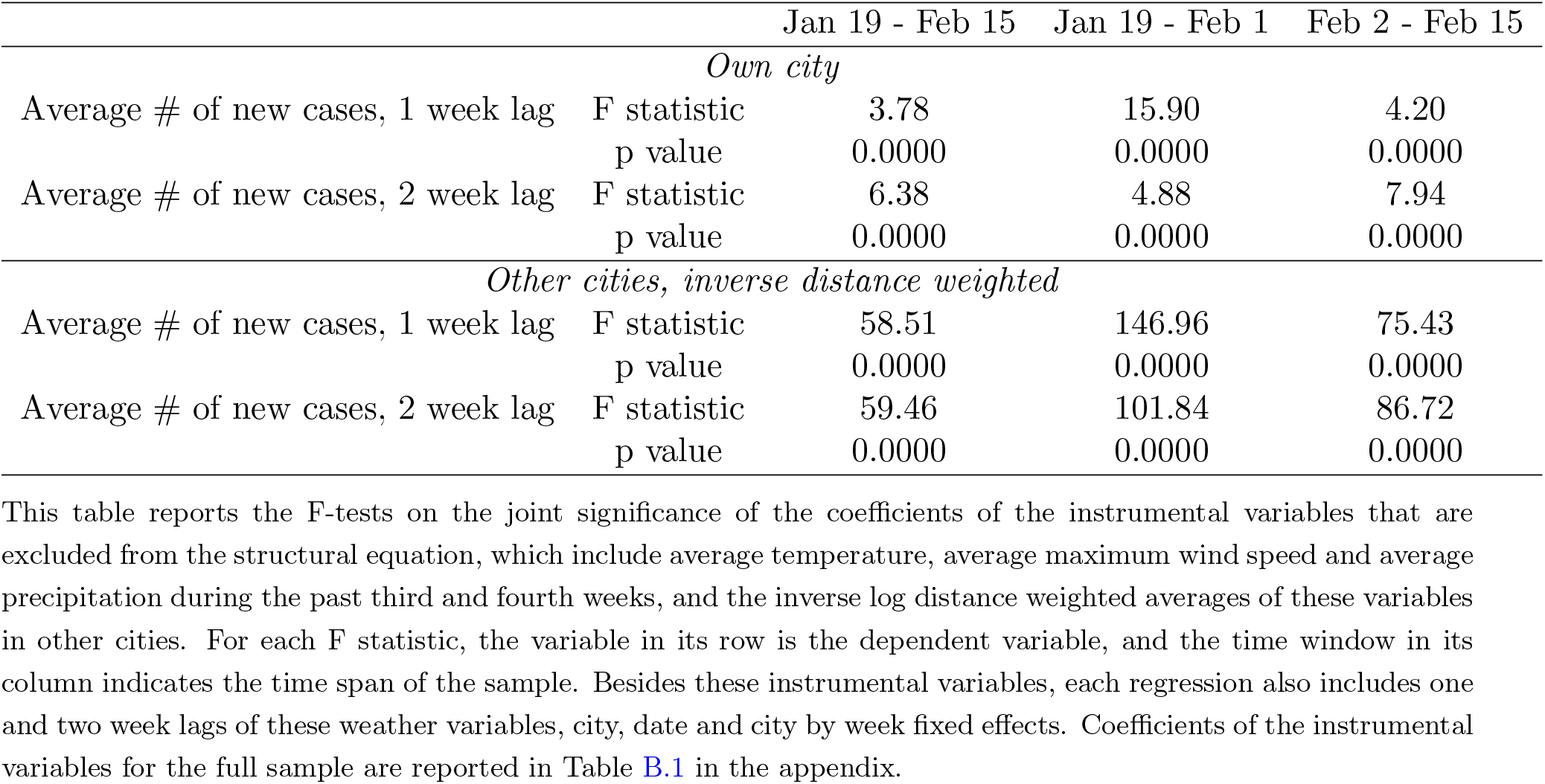
First Stage Results

|  |  | Jan 19 - Feb 15 | Jan 19 - Feb 1 | Feb 2 - Feb 15 |
| --- | --- | --- | --- | --- |
| *Own city* |  |  |  |  |
| Average # of new cases, 1 week lag | F statistic | 3.78 | 15.90 | 4.20 |
|  | p value | 0.0000 | 0.0000 | 0.0000 |
| Average # of new cases, 2 week lag | F statistic | 6.38 | 4.88 | 7.94 |
|  | p value | 0.0000 | 0.0000 | 0.0000 |
| *Other cities, inverse distance weighted* |  |  |  |  |
| Average # of new cases, 1 week lag | F statistic | 58.51 | 146.96 | 75.43 |
|  | p value | 0.0000 | 0.0000 | 0.0000 |
| Average # of new cases, 2 week lag | F statistic | 59.46 | 101.84 | 86.72 |
|  | p value | 0.0000 | 0.0000 | 0.0000 |

This table reports the F-tests on the joint significance of the coefficients of the instrumental variables that are excluded from the structural equation, which include average temperature, average maximum wind speed and average precipitation during the past third and fourth weeks, and the inverse log distance weighted averages of these variables in other cities. For each F statistic, the variable in its row is the dependent variable, and the time window in its column indicates the time span of the sample. Besides these instrumental variables, each regression also includes one and two week lags of these weather variables, city, date and city by week fixed effects. Coefficients of the instrumental variables for the full sample are reported in Table B.1 in the appendix.

## 4 Results

### 4.1 Within City Transmission

Our sample starts from January 19, when the first COVID-19 case was reported outside Wuhan. The sample spans four weeks in total and ends on February 15. We also estimate the model separately for each half sample (January 19 to February 1, and February 2 to February 15). The first two weeks witnessed the virus infections quickly spread throughout the country with every province reporting at least one confirmed case, and the number of cases also increased at increasing speed (Figure 1). During these two weeks, the government response has become more robust. On January 20, COVID-19 was classified as a Class B statutory infectious disease and treated as a Class A statutory infectious disease. The city of Wuhan was placed under lockdown on January 23, with road closures and residents not allowed to leave the city. Many other cities also imposed restrictions on travel, ranging from canceling public events, stopping public transportation to limiting how often residents can leave home. On February 8, the State Council issued The Notice on Orderly Resuming Work and Production in Enterprises, which is a defining event for the second phrase of the prevention and control measures in China (World Health Organization, 2020c). By comparing the dynamics of virus transmissions in these two half samples, we can evaluate the effectiveness of these public health measures.

Table 3 reports the estimation results of the OLS and IV regressions of Eq.(2), where only within city transmission is considered. After controlling for time-invariant city-specific effects and time effects common to all cities, on average, one new infection leads to 1.566 more cases in the next week, but 1.299 fewer cases one week later. The negative effect can be the result of both the local authorities and residents taking more protective measures in response to a higher perceived risk of contracting the disease. Releasing official information on newly confirmed cases at the daily level as well as exchanging information via social media throughout China may promote more timely actions to slowdown virus transmissions. Comparing containment effects over time, in the first half sample, one new infection leads to 1.966 more cases within a week, implying a fast growth in the number of cases. However, in the second half sample, the effect decreases to 0.490, suggesting that public health measures imposed in late January are effective in limiting a further spread of the virus.

**Table 3:** Within City Transmission of COVID-19

|  | Jan 19 - Feb 15 |  | Jan 19 - Feb 1 |  | Feb 2 - Feb 15 |  |
| --- | --- | --- | --- | --- | --- | --- |
|  | (1) | (2) | (3) | (4) | (5) | (6) |
|  | OLS | IV | OLS | IV | OLS | IV |
| *All Cities Excluding Wuhan* |  |  |  |  |  |  |
| Average # of new cases, 1 week lag | 0.834***<br>(0.00632) | 1.566***<br>(0.111) | 1.695***<br>(0.0376) | 1.966***<br>(0.0889) | -0.459***<br>(0.0598) | 0.490***<br>(0.0788) |
| Average # of new cases, 2 week lag | -0.389***<br>(0.00371) | -1.299***<br>(0.124) | 0.731<br>(2.007) | -1.678<br>(2.558) | -0.757***<br>(0.0310) | -0.847***<br>(0.0478) |
| Observations | 8,484 | 8,484 | 4,242 | 4,242 | 4,242 | 4,242 |
| Number of cities | 303 | 303 | 303 | 303 | 303 | 303 |
| Weather controls | YES | YES | YES | YES | YES | YES |
| City FE | YES | YES | YES | YES | YES | YES |
| Date FE | YES | YES | YES | YES | YES | YES |
| *All Cities Excluding Cities in Hubei Province* |  |  |  |  |  |  |
| Average # of new cases, 1 week lag | 0.745***<br>(0.0275) | 1.129***<br>(0.250) | 1.045***<br>(0.0827) | 1.345***<br>(0.194) | 0.0470<br>(0.126) | 0.344<br>(0.231) |
| Average # of new cases, 2 week lag | -0.472***<br>(0.0156) | -0.748***<br>(0.231) | 0.149<br>(0.683) | -1.751<br>(2.502) | -0.810***<br>(0.0746) | -0.758***<br>(0.188) |
| Observations | 8,036 | 8,036 | 4,018 | 4,018 | 4,018 | 4,018 |
| Number of cities | 287 | 287 | 287 | 287 | 287 | 287 |
| Weather controls | YES | YES | YES | YES | YES | YES |
| City FE | YES | YES | YES | YES | YES | YES |
| Date FE | YES | YES | YES | YES | YES | YES |

Besides Wuhan, a large number of cases are also reported in other cities in Hubei province, where 6 of them reported over 1000 cumulative cases by February 15^11^. Their overstretched health care system exacerbates the concern over delayed reporting of confirmed cases in these cities. To mitigate such potential measurement errors in our estimates, we exclude all cities in Hubei province from the regression. The bottom panel of Table 3 reports the estimates. Comparing the IV estimates in columns (4) and (6), we observe that in the first two weeks, on average, one new case leads to 1.345 more cases in the following week, while the chain of infections is broken in the second half sample.

### Between City Transmission

People may contract the virus via interacting with people who are infected and live in the same city or other cities. The severity of virus infections in other cities may influence the awareness of local public health authorities and residents. The rate of spread of the virus can be reduced if more protective measures are taken. In Eq.(1), we consider the effect of the infections in other cities and the epicenter of the epidemic, Wuhan, using inverse log distance as weights. The lockdown in Wuhan on January 23 significantly reduced the population flow from Wuhan to other cities, and geographic proximity may not describe well the level of social interactions between residents in Wuhan and other cities. To alleviate this concern, we use a measure of the size of population flow from Wuhan to a destination city, which is constructed by multiplying the daily migration index on the population flow out of Wuhan (Figure 2) with the share of the flow that a destination city receives provided by Baidu (Figure 3). During days before January 25, we use the average destination shares between January 10 and January 24. For days on or after January 24, the average destination shares between January 25 and February 23 are used^12^.

Table 4 reports the estimates from IV regressions of Eq.(1), and Table 5 reports the results where we exclude cities in Hubei province from the analysis. Column (4) of Table 4 indicates that in the first half sample, one new case leads to 1.465 more cases within one week, and the effect is not statistically significant between one and two weeks. In the second half sample, one new case actually decreases future number of new cases, which indicates that the responses by the government and the public effectively contain the risk of additional infections. The time varying patterns in local transmissions are evident using the rolling window analysis (Figure 4). The top left panel shows the estimated local transmission coefficients for various 14-day samples with the starting date indicated on the horizontal axis. After an initial increase in the local transmission rate, one case leads to fewer and fewer additional cases from late January onwards. Comparing Table 4 with Table 3, we observe that the results are not sensitive to the inclusion of terms on between-city transmissions.

**Table 4:** Within and Between City Transmission of COVID-19

|  | Jan 19 - Feb 15 |  | Jan 19 - Feb 1 |  | Feb 2 - Feb 15 |  |
| --- | --- | --- | --- | --- | --- | --- |
|  | (1) | (2) | (3) | (4) | (5) | (6) |
|  | OLS | IV | OLS | IV | OLS | IV |
| Average # of new cases, 1 week lag |  |  |  |  |  |  |
| Own city | 0.412***<br>(0.0276) | 0.804***<br>(0.0741) | 0.939***<br>(0.102) | 1.465***<br>(0.144) | -0.316***<br>(0.0614) | -0.0971***<br>(0.0258) |
| Other cities | -0.0143***<br>(0.00442) | -0.0755**<br>(0.0328) | 0.0895<br>(0.0717) | 0.0215<br>(0.0583) | -0.0483**<br>(0.0223) | -0.201***<br>(0.0628) |
| *weight = inverse distance* |  |  |  |  |  |  |
| Wuhan | -0.0695*<br>(0.0395) | 0.397<br>(0.310) | -0.852<br>(0.743) | -1.769*<br>(0.928) | -0.0294<br>(0.0243) | 0.118<br>(0.160) |
| *weight = inverse distance* |  |  |  |  |  |  |
| Wuhan | -0.00110**<br>(0.000493) | 0.00334***<br>(0.000575) | 0.00460***<br>(0.000342) | 0.00490***<br>(0.000470) | -0.00420***<br>(0.000711) | -0.00337***<br>(0.000727) |
| *weight = population flow* |  |  |  |  |  |  |
| Average # of new cases, 2 week lag |  |  |  |  |  |  |
| Own city | -0.259***<br>(0.0173) | -0.714***<br>(0.0594) | 2.547<br>(2.332) | 1.539<br>(2.502) | -0.434***<br>(0.0364) | -0.438***<br>(0.0755) |
| Other cities | 0.0477***<br>(0.0173) | 0.152***<br>(0.0483) | -0.358<br>(0.374) | 0.464<br>(0.397) | 0.0923**<br>(0.0390) | 0.456***<br>(0.158) |
| *weight = inverse distance* |  |  |  |  |  |  |
| Wuhan | 0.138<br>(0.0900) | -0.987<br>(0.707) | 2.883<br>(2.870) | 6.327*<br>(3.482) | 0.124*<br>(0.0715) | -0.916*<br>(0.519) |
| *weight = inverse distance* |  |  |  |  |  |  |
| Wuhan | 0.0134***<br>(0.000537) | 0.0116***<br>(0.000831) | 0.00714***<br>(0.00214) | 0.00146<br>(0.00224) | 0.00716***<br>(0.000384) | 0.00883***<br>(0.000476) |
| *weight = population flow* |  |  |  |  |  |  |
| Observations | 8,484 | 8,484 | 4,242 | 4,242 | 4,242 | 4,242 |
| Number of cities | 303 | 303 | 303 | 303 | 303 | 303 |
| Weather controls | YES | YES | YES | YES | YES | YES |
| City FE | YES | YES | YES | YES | YES | YES |
| Date FE | YES | YES | YES | YES | YES | YES |

**Table 5:** Within and Between City Transmission of COVID-19, Excluding Cities in Hubei Province

|  | Jan 19 - Feb 15 |  | Jan 19 - Feb 1 |  | Feb 2 - Feb 15 |  |
| --- | --- | --- | --- | --- | --- | --- |
|  | (1) | (2) | (3) | (4) | (5) | (6) |
|  | OLS | IV | OLS | IV | OLS | IV |
| Average # of new cases, 1 week lag |  |  |  |  |  |  |
| Own city | 0.617***<br>(0.0334) | 1.164***<br>(0.240) | 0.793***<br>(0.0827) | 1.517**<br>(0.661) | -0.0355<br>(0.149) | -0.0752<br>(0.191) |
| Other cities | 0.00266<br>(0.00183) | -0.0176<br>(0.0143) | -0.0167<br>(0.0199) | 0.0418<br>(0.179) | 3.30e-05<br>(0.00234) | -0.0429***<br>(0.0111) |
| *weight = inverse distance* |  |  |  |  |  |  |
| Wuhan | 0.00608<br>(0.0131) | 0.0629<br>(0.0513) | 0.160<br>(0.130) | 2.173*<br>(1.303) | 0.00785<br>(0.00752) | 0.110**<br>(0.0430) |
| *weight = inverse distance* |  |  |  |  |  |  |
| Wuhan | 0.00463***<br>(0.00147) | 0.00184<br>(0.00164) | 0.00636***<br>(0.00197) | 0.00105<br>(0.00350) | -0.0135**<br>(0.00643) | -0.00688<br>(0.00758) |
| *weight = population flow* |  |  |  |  |  |  |
| Average # of new cases, 2 week lag |  |  |  |  |  |  |
| Own city | -0.400***<br>(0.0264) | -0.724***<br>(0.142) | 0.256<br>(0.584) | 0.0862<br>(1.848) | -0.714***<br>(0.0435) | -0.724***<br>(0.0650) |
| Other cities | -0.000654<br>(0.00422) | 0.0409<br>(0.0309) | 0.183<br>(0.119) | -0.218<br>(1.473) | 0.00805<br>(0.00517) | 0.0728***<br>(0.0172) |
| *weight = inverse distance* |  |  |  |  |  |  |
| Wuhan | -0.0177<br>(0.0311) | -0.205<br>(0.150) | -0.777<br>(0.881) | -9.917*<br>(5.656) | -0.0432**<br>(0.0207) | -0.297***<br>(0.102) |
| *weight = inverse distance* |  |  |  |  |  |  |
| Wuhan | 0.00935***<br>(0.00165) | 0.00246<br>(0.00394) | 0.00974***<br>(0.00328) | -0.00476<br>(0.0106) | 0.00352***<br>(0.00118) | 0.00645***<br>(0.00150) |
| *weight = population flow* |  |  |  |  |  |  |
| Observations | 8,036 | 8,036 | 4,018 | 4,018 | 4,018 | 4,018 |
| Number of cities | 287 | 287 | 287 | 287 | 287 | 287 |
| Weather controls | YES | YES | YES | YES | YES | YES |
| City FE | YES | YES | YES | YES | YES | YES |
| Date FE | YES | YES | YES | YES | YES | YES |

**Figure 4:**
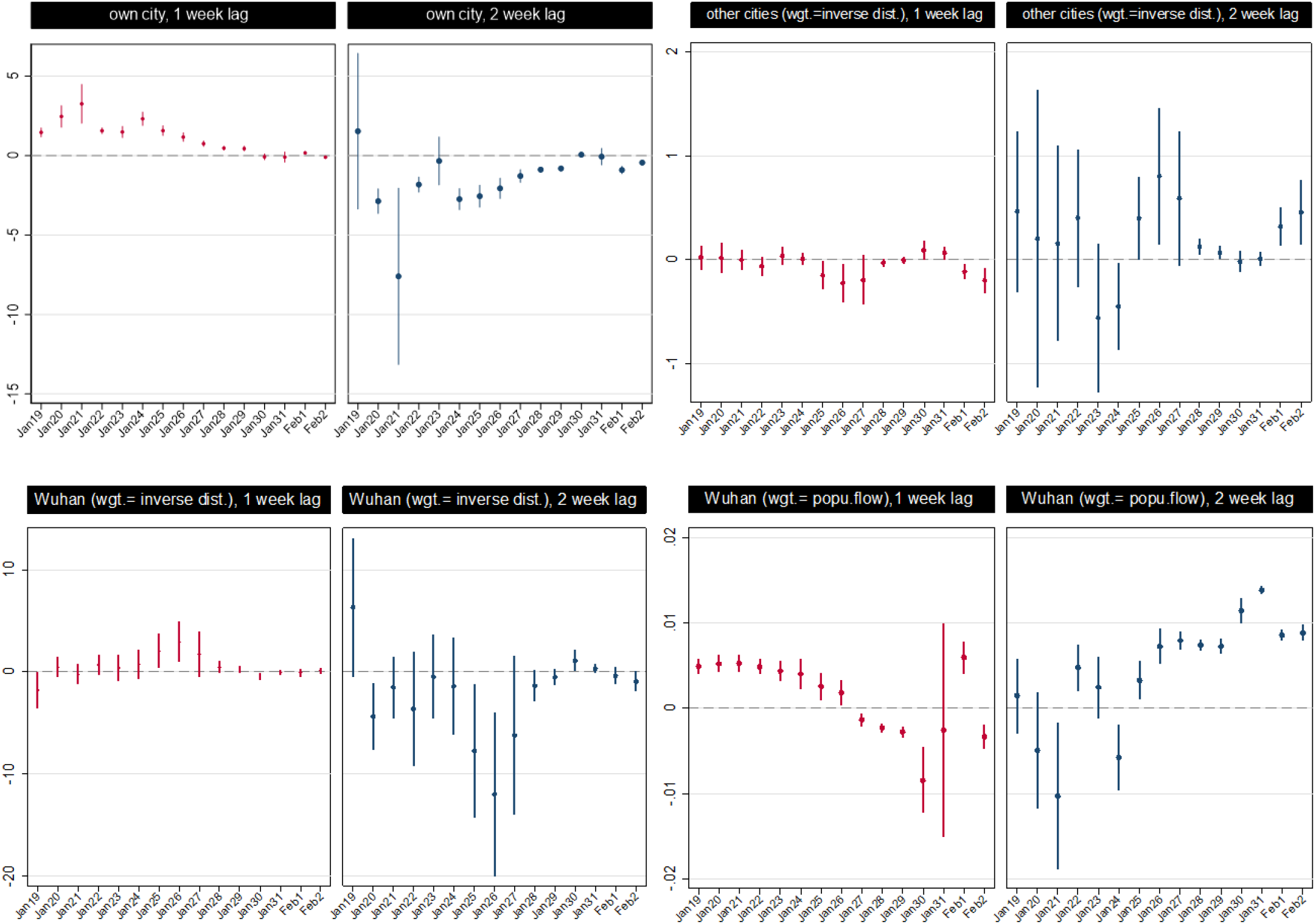
Rolling Window Analysis of Within and Between City Transmission of COVID-19 This table shows the estimated coefficients and 95% CIs from instrumental variable regressions of daily number of new confirmed COVID-19 cases on average numbers of new cases in the preceding one and two weeks, in the same city, nearby cities (weighted by inverse log distance) and Wuhan (weighted by inverse log distance or population flow). Each estimation sample contains 14 days with the starting date indicated on the horizontal axis.

As a robustness test, Table 5 reports the estimation results if the analysis sample does not include cities in Hubei province. Column (4) of Table 5 indicates that in the first half sample, one new case leads to 1.517 more cases within a week, and this becomes small and statistically not significant in the second half sample. Besides, in the second half sample, one new case decreases new infections by 0.724 between 1 and 2 weeks, which is larger than the estimate (0.438) with cities in Hubei province included. Overall, the spread of the virus has been effectively contained between February 2 and February 15, particularly for cities outside Hubei province.

In the epidemiology literature, the estimates on the basic reproduction number of COVID-19 is approximately within the range 2 *∼* 2.5 (World Health Organization, 2020c). Its value depends on the model used and factors that affect disease transmissions such as the behavior of the susceptible and infected population. Intuitively, it can be interpreted as measuring the expected number of new cases that are generated by one existing case. It is of interest to note that our estimates are at the lower end of the range. One more case leads to 1.465 more cases in the same city in the next week (1.517 if cities in Hubei province are excluded). The effects on further lags or in the second half sample (February 2 - February 15) are small, negative, or not precisely estimated, suggesting that factors such as public health measures and people’s behavior may play an important role in containing the transmission of COVID-19.

For the between-city transmission from Wuhan, we observe that the population flow better explains the contagion effect than geographic proximity (Table 4). In the first half sample, one new case in Wuhan leads to more cases in other cities which receive more population flows from Wuhan within one week. In the second half sample, more arrivals from Wuhan one week earlier can still be a risk. A back of the envelope calculation indicates that one new case in Wuhan leads to 0.067 (0.120) more cases in the destination city per 10,000 travelers from Wuhan within one (two) week between January 19 and February 1 (February 2 and February 15)^13^. Note that while the effect is statistically significant, it should be interpreted in context. It was estimated that 15, 000, 000 people would travel out of Wuhan during the Lunar New Year holiday^14^. If all had gone to one city, this would have directly generated about 280.5 cases within two weeks. The risk of infection is likely very low for most travelers except for few who have previous contacts with sources of infection, and person-specific history of past contacts may be an essential predictor for infection risk, in addition to the total number of population flows^15^.

Besides spillovers from Wuhan, a city may also be affected by infections nearby. We observe that in the second half sample, one new case in a neighboring city that is 100km away generates 0.099 more cases after between one and two weeks^16^. This indicates that by February, while within city transmissions are effectively contained, new infections caused by between-city transmissions can still be a risk. For cities outside Hubei province, the results are similar (Table 5), except that the transmission from Wuhan is not significant in the first half sample.

### 4.3 Social and Economic Mediating Factors

We investigate the impacts of many social, economic and environmental variables in mediating the transmission rates (Eq.(3)). For own city transmissions, we consider the mediating effects of population density, per capita GDP, number of doctors, contemporaneous temperature, wind speed and precipitation. For between-city transmissions, we consider the mediating effects of distance, density difference and per capita GDP difference. We also include a measure of population flows from Wuhan. Table 6 reports the estimation results of the IV regressions using the first half sample, and Table 7 reports the estimates for the second half sample.

**Table 6:** Social and Economic Factors Mediating the Transmission of COVID-19, Jan 19 - Feb 1

|  | (1) | (2) | (3) | (4) | (5) |
| --- | --- | --- | --- | --- | --- |
| Average # of new cases, 1 week lag |  |  |  |  |  |
| Own city | 1.290<br>(0.984) | 0.741<br>(0.631) | 0.511<br>(0.658) | 0.435<br>(0.656) | 0.0292<br>(0.651) |
| × population density | -0.000735<br>(0.000518) |  | -0.000890***<br>(0.000283) | -0.00104**<br>(0.000406) | -0.00105***<br>(0.000382) |
| × per capita GDP | 0.154<br>(0.122) |  | 0.237**<br>(0.112) | 0.230**<br>(0.114) | 0.290***<br>(0.110) |
| × # of doctors | -0.179*<br>(0.0989) |  | -0.0293<br>(0.141) | -0.0124<br>(0.134) | 0.00240<br>(0.143) |
| × temperature |  | -0.0781<br>(0.0531) | -0.0173<br>(0.0705) | -0.0186<br>(0.0732) | -0.0307<br>(0.0772) |
| × wind speed |  | 0.151<br>(0.223) | -0.0653<br>(0.0819) | -0.0341<br>(0.0770) | -0.0479<br>(0.0784) |
| × precipitation |  | -0.575<br>(0.440) | 0.0858<br>(0.294) | 0.102<br>(0.278) | 0.147<br>(0.307) |
| Other cities | 0.0588<br>(0.0941) | -0.0739<br>(0.0930) | -0.0123<br>(0.0527) | 0.00975<br>(0.0478) | 0.00741<br>(0.0608) |
| *weight = inverse distance* |  |  |  |  |  |
| Other cities |  |  |  | -0.00619<br>(0.00467) | 0.00132<br>(0.00423) |
| *weight = inverse density ratio* |  |  |  |  |  |
| Wuhan | -3.518***<br>(0.608) | -2.192<br>(1.744) | -2.537**<br>(1.241) | -2.540**<br>(1.256) | -2.190<br>(1.363) |
| *weight = inverse distance* |  |  |  |  |  |
| Wuhan | 0.00639***<br>(0.000672) | 0.00564***<br>(0.000895) | 0.00597***<br>(0.000690) | 0.00589***<br>(0.000707) | 0.00601***<br>(0.000678) |
| *weight = population flow* |  |  |  |  |  |
| Wuhan |  |  |  |  | -0.0562<br>(0.766) |
| *weight = inverse per capita GDP ratio* |  |  |  |  |  |
| Wuhan |  |  |  |  | 0.0270<br>(0.0238) |
| *weight = inverse density ratio* |  |  |  |  |  |
| Average # of new cases, 2 week lag |  |  |  |  |  |
| Own city | 16.93*<br>(10.25) | 36.28***<br>(12.46) | 45.21**<br>(19.15) | 43.77**<br>(18.95) | 45.87**<br>(18.65) |
| × population density | 0.00343<br>(0.00335) |  | 0.00663***<br>(0.00252) | 0.00706**<br>(0.00281) | 0.00696***<br>(0.00248) |
| × per capita GDP | -1.106<br>(1.277) |  | -0.151<br>(0.639) | -0.174<br>(0.609) | -0.0853<br>(0.643) |
| × # of doctors | -1.094<br>(0.987) |  | -4.050**<br>(1.897) | -3.941**<br>(1.758) | -4.323**<br>(1.958) |
| × temperature |  | -0.296<br>(0.529) | -1.429**<br>(0.592) | -1.379**<br>(0.565) | -1.427**<br>(0.594) |
| × wind speed |  | -7.252*** | -3.577 | -3.557 | -3.657 |
| Continued on next page |  |  |  |  |  |

| Table 6 – continued from previous page | (1) | (2) | (3) | (4) | (5) |
| --- | --- | --- | --- | --- | --- |
|  |  | (1.954) | (2.547) | (2.558) | (2.784) |
| × precipitation |  | -2.604 | -23.55*** | -22.18*** | -20.14*** |
|  |  | (10.32) | (5.654) | (5.412) | (4.719) |
| Other cities | -0.188 | 1.415** | 0.305 | 0.156 | 0.138 |
| *weight = inverse distance* | (0.926) | (0.614) | (0.476) | (0.458) | (0.543) |
| Other cities |  |  |  | 0.0505 | -0.0615 |
| *weight = inverse density ratio* |  |  |  | (0.0530) | (0.0582) |
| Wuhan | 14.21*** | 8.903 | 11.02** | 10.93** | 9.217* |
| *weight = inverse distance* | (2.538) | (7.749) | (5.492) | (5.194) | (5.553) |
| Wuhan | -0.000999 | 0.00236 | 0.00421 | 0.00479 | 0.00718 |
| *weight = population flow* | (0.00408) | (0.00205) | (0.00490) | (0.00492) | (0.00468) |
| Wuhan |  |  |  |  | -3.547 |
| *weight = inverse per capita GDP ratio* |  |  |  |  | (3.434) |
| Wuhan |  |  |  |  | -0.0279 |
| *weight = inverse density ratio* |  |  |  |  | (0.113) |
| Observations | 4,242 | 4,242 | 4,242 | 4,242 | 4,242 |
| Number of cities | 303 | 303 | 303 | 303 | 303 |
| Weather controls | YES | YES | YES | YES | YES |
| City FE | YES | YES | YES | YES | YES |
| Date FE | YES | YES | YES | YES | YES |

**Table 7:** Social and Economic Factors Mediating the Transmission of COVID-19, Feb 2 - Feb 15

|  | (1) | (2) | (3) | (4) | (5) |
| --- | --- | --- | --- | --- | --- |
| Average # of new cases, 1 week lag |  |  |  |  |  |
| Own city | -0.127 | 0.745* | -0.683 | -0.684 | -0.672 |
|  | (0.126) | (0.407) | (0.718) | (0.633) | (0.662) |
| × population density | -0.000171 |  | 0.000712* | 0.000596 | 0.000609 |
|  | (0.000222) |  | (0.000416) | (0.000396) | (0.000490) |
| × per capita GDP | 0.105*** |  | 0.252*** | 0.247*** | 0.240*** |
|  | (0.0201) |  | (0.0624) | (0.0620) | (0.0544) |
| × # of doctors | -0.195* |  | -0.589*** | -0.548*** | -0.537*** |
|  | (0.110) |  | (0.227) | (0.210) | (0.208) |
| × temperature |  | -0.130*** | 0.0220 | 0.0196 | 0.0167 |
|  |  | (0.0242) | (0.0756) | (0.0668) | (0.0689) |
| × wind speed |  | 0.0432 | -0.148* | -0.122 | -0.115 |
|  |  | (0.0865) | (0.0874) | (0.0969) | (0.0974) |

| **Table 7 – continued from previous page** | (1) | (2) | (3) | (4) | (5) |
| --- | --- | --- | --- | --- | --- |
| × precipitation |  | 1.748* | 2.192* | 2.037* | 1.869* |
|  |  | (0.915) | (1.177) | (1.182) | (1.125) |
| Other cities | -0.182*** | -0.0825*** | -0.146 | -0.125 | -0.128 |
| *weight = inverse distance* | (0.0493) | (0.0271) | (0.111) | (0.0921) | (0.0907) |
| Other cities |  |  |  | -0.000483 | -0.00330 |
| *weight = inverse density ratio* |  |  |  | (0.00230) | (0.00504) |
| Wuhan | 0.0228 | 0.262 | 0.360 | 0.234 | 0.342 |
| *weight = inverse distance* | (0.165) | (0.300) | (0.417) | (0.315) | (0.404) |
| Wuhan | -0.00334** | -0.0138*** | -0.0112*** | -0.0107*** | -0.0113*** |
| *weight = population flow* | (0.00134) | (0.00398) | (0.00413) | (0.00388) | (0.00404) |
| Wuhan |  |  |  |  | 0.102 |
| *weight = inverse per capita GDP ratio* |  |  |  |  | (0.268) |
| Wuhan |  |  |  |  | -0.00457 |
| *weight = inverse density ratio* |  |  |  |  | (0.00923) |
| Average # of new cases, 2 week lag |  |  |  |  |  |
| Own city | -0.351 | -1.163 | -0.819 | -0.862 | -0.774 |
|  | (0.234) | (0.858) | (1.028) | (1.003) | (0.916) |
| × population density | -0.000442*** |  | 0.000286 | 0.000181 | 0.000289 |
|  | (0.000148) |  | (0.000302) | (0.000337) | (0.000342) |
| × per capita GDP | 0.0631*** |  | 0.165* | 0.163* | 0.131 |
|  | (0.0189) |  | (0.0874) | (0.0867) | (0.0859) |
| × # of doctors | -0.0364 |  | -0.223*** | -0.209*** | -0.201** |
|  | (0.0709) |  | (0.0763) | (0.0748) | (0.0863) |
| × temperature |  | 0.222* | 0.166 | 0.165 | 0.162 |
|  |  | (0.122) | (0.134) | (0.129) | (0.120) |
| × wind speed |  | -0.195** | -0.375** | -0.346** | -0.332** |
|  |  | (0.0836) | (0.150) | (0.153) | (0.160) |
| × precipitation |  | 1.671*** | 1.619 | 1.462 | 1.362 |
|  |  | (0.647) | (1.295) | (1.320) | (1.249) |
| Other cities | 0.364*** | 0.198*** | 0.213 | 0.194* | 0.208* |
| *weight = inverse distance* | (0.120) | (0.0616) | (0.134) | (0.115) | (0.120) |
| Other cities |  |  |  | 0.00181 | 0.000865 |
| *weight = inverse density ratio* |  |  |  | (0.00390) | (0.00263) |
| Wuhan | -0.487* | -0.634 | -0.963 | -0.713 | -0.961 |
| *weight = inverse distance* | (0.285) | (0.666) | (1.003) | (0.803) | (1.018) |
| Wuhan | 0.00888*** | 0.0148*** | 0.0132*** | 0.0129*** | 0.0126*** |
| *weight = population flow* | (0.000398) | (0.00151) | (0.000774) | (0.000742) | (0.000721) |
| Wuhan |  |  |  |  | -0.298 |
| *weight = inverse per capita GDP ratio* |  |  |  |  | (0.834) |
| Wuhan |  |  |  |  | 0.0167 |
| *weight = inverse density ratio* |  |  |  |  | (0.0241) |

| **Table 7 – continued from previous page** | (1) | (2) | (3) | (4) | (5) |
| --- | --- | --- | --- | --- | --- |
| Observations | 4,242 | 4,242 | 4,242 | 4,242 | 4,242 |
| Number of cities | 303 | 303 | 303 | 303 | 303 |
| Weather controls | YES | YES | YES | YES | YES |
| City FE | YES | YES | YES | YES | YES |
| Date FE | YES | YES | YES | YES | YES |

Our main conclusions still hold with these mediating factors included. The effects of past infections are much smaller in the second half sample, suggesting that the virus outbreak has been contained. For the between-city transmissions, population flow from Wuhan poses a risk for new infections in other cities, with one week lag in the first half sample, and with two week lag in the second half sample. The effects are robust with the inclusion of other measures of proximity, such as those based on geography and economic similarity.

Table 8 shows variables that affect transmissions within cities and whose coefficients are statistically significant at 0.1. In order to help compare across variables, we compute the changes in the variables needed in order for the coefficient of past infections on current cases to be reduced by 1. The effects of the environmental factors and population density are mixed. We also observe that the transmission rate is lower in cities with more doctors. Cities with higher per capita GDP have higher transmission rates, which may be due to more social interactions as a result of more economic activity^17^. However, until clearer mechanisms are identified and tested, this should be interpreted with caution.

**Table 8:** A Summary of Factors Mediating Within City Transmissions of COVID-19

| for local transmission rate to be reduced by 1 |  | Jan 19 - Feb 1 | Feb 2 - Feb 15 |
| --- | --- | --- | --- |
| lag 1 week | precipitation |  | −0.54 mm |
|  | population density | +952.38 per km<sup>2</sup> |  |
|  | # of doctors |  | +18,622 |
|  | per capita GDP | −34,483 RMB | −41,667 RMB |
| lag 2 week | wind speed |  | +3.01 m/s |
|  | temperature | +0.70C |  |
|  | precipitation | +0.05 mm |  |
|  | population density | −143.68 per km<sup>2</sup> |  |
|  | # of doctors | +2,313 | +49,751 |

This table shows changes in the variables needed in order for the impact of past same city infections on current infections to be reduced by 1, using estimates in Column (5) of Tables 6 and 7, with significance levels of at least 0.1.

## 5 Conclusion

This paper examines the transmission dynamics of the novel coronavirus 2019, considering both within- and between-city transmissions. Changes in weather conditions induce exogenous variations in past infection rates, which allows us to identify the causal impact of past infections on new cases. We use a machine learning method (Lasso) to select instrumental variables with strong predictive power for the endogenous variables. The estimates show that the positive effect of an infection in generating new local infections is observed within one week, and there is evidence that people’s responses can break the chain of infections. Comparing estimates in two half samples, we observe that the spread of COVID-19 has been effectively contained by early February, especially for cities outside Hubei province, the epicenter of the outbreak. Data on real-time population flows between cities have become available in recent years, after aggregating location data from various sources of individual mobile phone users. We show that this new source of data is valuable in explaining between-city transmissions of COVID-19, even after controlling for traditional measures of geographic and economic proximity.

At the time of writing, COVID-19 represents a very high risk at the global level, according to the WHO risk assessment (World Health Organization, 2020a). Countries other than China are reporting new confirmed cases, and increasingly many cases arise through community transmissions rather than being imported. Based on case data in China between January 19 and February 15, we show that public health measures adopted can effectively contain the virus outbreak, and newly available data on real-time population flows can be a valuable tool for risk assessment by public health authorities and the general public.

A key limitation of the paper is that we are not able to disentangle the effects from each of the stringent measures taken, as even within this four-week sampling period China enforced such a large number of densely timed policies to contain the virus spreading, often simultaneously in many cities. A second limitation is that these policies fall within each of the two sub-samples. By the starting date of the official data release for confirmed infected cases throughout China, i.e. January 2020, a number of stringent measures were already taken, which prevent researchers to ideally examine and compare a sub-sample period during which no strict policies was enforced. Key knowledge gaps remain in the understanding of the epidemiological characteristics of COVID-19, such as individual risk factors for contracting the virus and infections from asymptotic cases. Data on the demographics and exposure history for those who have shown symptoms as well as those who have not will help facilitate these research.

## Data Availability

Data on the daily number of newly confirmed COVID-19 cases at the city level in China are collected from the websites of provincial health commissions of China. Data on the daily weather characteristics are collected from the NOAA website. Data on the daily population flow intensity index from Wuhan to other cities are collected from the Baidu website. All these data are publicly available. The city-level characteristics are collected from China City Statistical Yearbooks.

https://www.ncdc.noaa.gov/cdo-web/

http://qianxi.baidu.com/?from=groupmessage

## A Data

Figure A1 illustrates the dependent variable, the instrumental variables, and the exclusion restriction that weather conditions earlier than two weeks from date *t* affect the number of people who are infectious within past two weeks, but do not directly affect the number of confirmed new cases at date *t*.

**Figure A.1:**
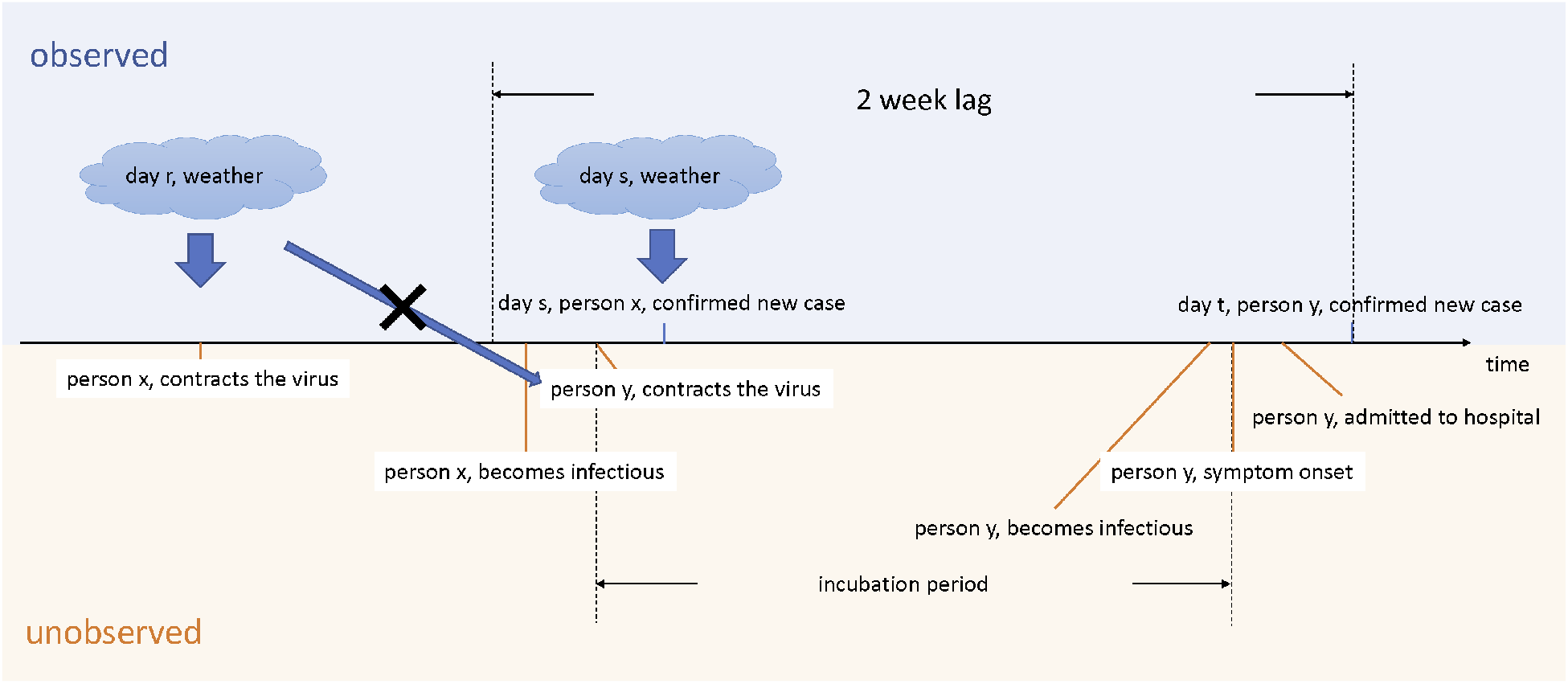
Timeline of Key Variables

## B First Stage Regressions

Table B.1 reports results of the first stage of the IV regressions (Table 4). In Columns (1) and (2), the dependent variables are the numbers of newly confirmed COVID-19 cases in the own city in the preceding first and second weeks, respectively. In Columns (3) and (4), the dependent variables are the weighted average numbers of newly confirmed COVID-19 cases in other cities in the preceding first and second weeks (weighted by the inverse log distance), respectively. The weather variables in the preceding first and second weeks are included in the control variables. The weather variables in the preceding third and fourth weeks are the excluded instruments in the IV regressions, and their coefficients are reported in the table.

Weather conditions affect disease transmissions either directly if the virus can more easily survive and spread in certain environment, or indirectly by changing human behavior. In our sample period, higher wind speeds or more precipitation decrease the number of infections in the future. Higher temperatures increase local infections, but decrease infections in other cities.

**Table B.1:** First Stage Regressions

| Dependent variable | Average # of new cases |  |  |  |
| --- | --- | --- | --- | --- |
|  | Own city |  | Other cities |  |
|  | 1 week lag<br>(1) | 2 week lag<br>(2) | 1 week lag<br>(3) | 2 week lag<br>(4) |
| *Own city* |  |  |  |  |
| Weekly average temperature, 3 week lag | 0.194<br>(1.080) | -0.163<br>(0.410) | -2.542***<br>(0.940) | -2.597***<br>(0.477) |
| Weekly average maximum wind speed, 3 week lag | 0.582<br>(2.066) | -0.177<br>(0.785) | -0.712<br>(1.798) | 0.131<br>(0.912) |
| Weekly average precipitation, 3 week lag | -0.305<br>(4.304) | 0.180<br>(1.636) | -5.022<br>(3.745) | -4.309**<br>(1.900) |
| Weekly average temperature, 4 week lag | 0.650<br>(0.894) | 0.390<br>(0.340) | 0.436<br>(0.778) | -0.0727<br>(0.395) |
| Weekly average maximum wind speed, 4 week lag | 0.767<br>(1.568) | 0.0529<br>(0.596) | -0.828<br>(1.364) | -0.247<br>(0.692) |
| Weekly average precipitation, 4 week lag | -6.467<br>(3.982) | -3.572**<br>(1.514) | -3.852<br>(3.465) | -3.780**<br>(1.758) |
| *Other cities, weight = inverse distance* |  |  |  |  |
| Weekly average temperature, 3 week lag | 0.592<br>(0.469) | 0.385**<br>(0.178) | 2.832***<br>(0.408) | 2.036***<br>(0.207) |
| Weekly average maximum wind speed, 3 week lag | -3.624***<br>(1.041) | -1.725***<br>(0.396) | -12.49***<br>(0.906) | -6.751***<br>(0.460) |
| Weekly average precipitation, 3 week lag | -4.574**<br>(2.210) | -2.019**<br>(0.840) | -16.40***<br>(1.923) | -6.457***<br>(0.976) |
| Weekly average temperature, 4 week lag | 0.283<br>(0.390) | 0.184<br>(0.148) | 1.366***<br>(0.339) | 1.050***<br>(0.172) |
| Weekly average maximum wind speed, 4 week lag | -2.102**<br>(0.822) | -1.047***<br>(0.313) | -6.425***<br>(0.715) | -3.681***<br>(0.363) |
| Weekly average precipitation, 4 week lag | -0.0551<br>(1.856) | -0.0585<br>(0.705) | -6.355***<br>(1.615) | -3.597***<br>(0.819) |
| F statistic | 3.78 | 6.38 | 58.51 | 59.46 |
| p value | 0.0000 | 0.0000 | 0.0000 | 0.0000 |
| Observations | 8,512 | 8,512 | 8,512 | 8,512 |
| Number of cities | 304 | 304 | 304 | 304 |
| Weather controls | YES | YES | YES | YES |
| City FE | YES | YES | YES | YES |
| Date FE | YES | YES | YES | YES |
| City by Week FE | YES | YES | YES | YES |

This table shows the results of the first stage IV regressions. Coefficients of the weather variables which are used as excluded instrumental variables are reported. \*\*\* p<0.01, \*\* p<0.05, \* p<0.1.

## Footnotes

1 COVID-19 is previously known as novel coronavirus pneumonia or 2019-nCoV acute respiratory disease.

2 https://gisanddata.maps.arcgis.com/apps/opsdashboard/index.html#/bda7594740fd40299423467b48e9ecf6

3 According to Law of the People’s Republic of China on Prevention and Treatment of Infectious Diseases, Class A infectious diseases only include plague and cholera.

4 Li *et al*. (2020) document the exposure history of the first 425 cases. It is suspected that the initial cases were linked to the Huanan Seafood Wholesale Market in Wuhan.

5 The 2019-nCoV epidemic is still ongoing and the estimates are revised from time to time in the literature as new data become available. The current estimates include the following. The incubation period is estimated to be between 2 and 10 days (World Health Organization, 2020b), 5.2 days (Li *et al*. (2020)), or 3 days (median, Guan *et al*. (2020)). The average infectious period is estimated to be 1.4 days (Wu *et al*., 2020a).

6 On February 12, cities in Hubei province include clinically diagnosed cases in the confirmed cases, in addition to cases that are confirmed by nucleic acid tests, which results in a sharp increase in the number of confirmed cases for cities in Hubei on February 12. The common effect on other cities is controlled for by the day fixed effect.

7 Flu viruses are easier to survive in cold weather. Adverse weather conditions also limit outdoor activities which can decrease the chance of contracting the virus. For details, see Adda (2016) and Section 3.2.

8 Hong Kong and Macao are excluded from our analysis due to the lack of some socioeconomic variables.

9 Baidu Migration, qianxi.baidu.com.

10 The 100km circle is consistent with the existing literature. Most studies on the socioeconomic impacts of climate change have found that estimation results are insensitive to the choice of the cutoff distance (Zhang *et al*., 2017). We also used other cutoff distances (e.g., 150km) for robustness checks, and the main results remain unchanged. These results are available upon request.

11 These cities are Xiaogan, Huanggang, Jingzhou, Suizhou, Ezhou, and Xiangyang.

12 The shares of top 100 destinations are available. The starting and ending dates of the average shares released by Baidu do not precisely match the period of the analysis sample.

13 It is estimated that 14,925,000 people traveled out of Wuhan in 2019 during the Lunar New Year holiday (http://www.whtv.com.cn/p/17571.html). The sum of Baidu’s migration index for population flow out of Wuhan during the 40 days around the 2019 Lunar New Year is 203.3, which means one index unit represents 0.000013621 travelers. The destination share is in percentage. With one more case in Wuhan, the effect on a city receiving 10,000 travelers from Wuhan is 0.00490 *×* 0.000013621 *×* 100 *×* 10000 = 0.067.

14 http://www.whtv.com.cn/p/17571.html

15 From mid February, individual specific health codes such as Alipay Health Code and WeChat Health Code are being used in many cities to aid quarantine efforts.

16 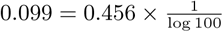

17 Fogli and Veldkamp (2019) show that income is positively correlated with more closely connected social network, and a dense network spreads diseases faster.

